# Variation in bulk RNA-seq and estimated cell type proportion using deconvolution when comparing pancreatic cancer samples within the same individual

**DOI:** 10.1101/2025.05.05.25326976

**Authors:** Rick J. Jansen, Sarah A. Munro, Samuel O. Antwi, Kari G. Rabe, Hugues Sicotte

## Abstract

**Introduction:** There is great promise in using genomic data to inform individual cancer treatment plans. Assessing intratumor genetic heterogeneity, studies have shown it may be possible to target biopsies to tumor subclones driving disease progression or treatment resistance. Here, we explore if the interpretation of tumor gene expression analysis varies across two specimens from the same patient. **Methods:** We performed bulk RNA-seq using FFPE samples from 16 patients who also had a previous separate bulk RNA-seq performed and deposited in TCGA. We used three different deconvolution methods to compare cell type proportions for these paired data. We normalized study-specific gene expression values per gene by calculating transcripts per million and adjusted for batch effect across study to compare median expression values. We also compared the reliability of gene expression measurements. We selected *KRAS, TP53, SMAD4*, and *CDKN2A*, as the most mutated genes in pancreatic cancer, and *CTNNB1, JUN, SMAD3, SMAD7*, and *TCF7,* as these tend to be enriched in pancreatic cancer compared with adjacent normal tissue. **Results:** We found that average cell type proportion varied the most between studies (i.e., samples for each patient) for NK and macrophages (using adjusted p-value 0.05/21=0.002). For the differential expression analysis, we did not observe significant differences in average expression of any of the selected genes. We observed substantial (kappa=0.75) for only *JUN* with low to moderate concordance (i.e., Kappa value 0.25-0.5) when using a median cut point for the remaining 8 genes across the two studies. **Discussion:** Together, the findings suggest that more than one tumor sample may be needed for effective treatment planning. Any potential difference in observed expression values across the paired samples could be related to the different cell type proportions across the samples. The sample size was small, and each study used different sequencing technologies, so any interpretation should be confirmed with additional studies.

## Introduction

Pancreatic Ductal Adenocarcinoma (PDAC) is a highly aggressive malignancy with heterogeneous tumor microenvironment.(Oketch et al., 2024) As a result, patients are often diagnosed at a late stage contributing to its high mortality rate and poor treatment response.(Grünwald et al., 2021; Hwang et al., 2022) In 2020, approximately 495,773 new cases were diagnosed worldwide, ranking it as the 12th most common malignancy.(Cai et al., 2021; J. X. Hu et al., 2021; Li et al., 2024; Rawla et al., 2019) In the United States, the American Cancer Society reported approximately 60,430 new cases and 48,220 deaths in 2021, ranking pancreatic cancer as the third leading cause of cancer death.(Cai et al., 2021; J. X. Hu et al., 2021) Similarly, in the European Union, it is projected that approximately 111,500 people will die from pancreatic cancer by 2025. These statistics underscore the urgent need for improved prevention, early detection, and treatment strategies to mitigate the escalating impact of pancreatic cancer on global health.(Ying et al., 2025)

The promise and utility of using genomic data to inform individual cancer treatment plans has been longstanding. However, minimal attention has been focused on using genomic data to inform biopsy protocols or guide biopsy sampling strategies to enhance the diagnostic and prognostic yield of tissue sampling. By assessing intratumor genetic heterogeneity, studies in other cancers have shown it may be possible to target biopsies to tumor subclones driving disease progression or treatment resistance.(Blanco-Heredia et al., 2024) The tumor microenvironment (TME) in PDAC is a complex biological barrier with multiple components, such as desmoplasia, hypoxia, presence of various cell types, and complex signaling pathways, making treatment challenging.(Binkowski et al., 2024, 2025; Giannoukakos et al., 2024; Padwal et al., 2024)

The TME’s complexity necessitates innovative therapeutic strategies, and nanomedicine offers potential solutions for targeted drug delivery and modulation of the microenvironment.(Nair et al., 2024) CNV studies analyze the expression levels of genes with amplifications or deletions in malignant tissues compared to normal pancreatic tissues to understand the CNV landscape and identify correlations with survival outcomes.(Giannoukakos et al., 2024) These variations can influence gene expression, leading to the dysregulation of critical cellular processes and contributing to cancer development and progression.(Mazur et al., 2010) Liquid biopsy, particularly RNA sequencing (RNA-Seq) of blood samples, is emerging as a non-invasive method for cancer diagnostics and monitoring treatment response.(Giannoukakos et al., 2024) RNA-seq allows for the analysis of gene expression and identification of potential biomarkers in body fluids.(Padwal et al., 2024) Studies employing RNA-Seq have identified potential biomarkers for early detection of pancreatic cancer, including with proteins.(Butera et al., 2024; Karmakar et al., 2020; R. Liu et al., 2024) Furthermore, some research indicates that a NETseq ensemble classifier based on peripheral blood RNA-seq data may serve as a tool for non-invasive detection of neuroendocrine tumors and assessment of treatment response.(Padwal et al., 2024)

Here, we explored the importance of targeted biopsy sampling by using bulk RNA-seq data to analyze variation gene expression across paired tumor samples from the same group of individuals.

## Materials and Methods

We performed bulk RNA-seq extracted from FFPE samples of 16 patients who also had bulk RNA-seq performed on a different tumor section and deposited in TCGA. The pipeline and workflow details for the TCGA can be found on their website. (https://docs.gdc.cancer.gov/Data/Bioinformatics_Pipelines/Expression_mRNA_Pipeline/) The second sample for these16 patients was processed using the NovaSeq S4 PE100 with Illumina’s TruSeq Total Stranded RNA prep reagents (https://www.illumina.com/products/by-type/sequencing-kits/library-prep-kits/truseq-stranded-total-rna.html) and used a second DNase treatment to minimize any potential DNA contamination. Table 1 shows select patient characteristics for our study. (**Table 1**) The Institutional Review Board of Mayo Clinic gave approval for this work.

**Table 1.**
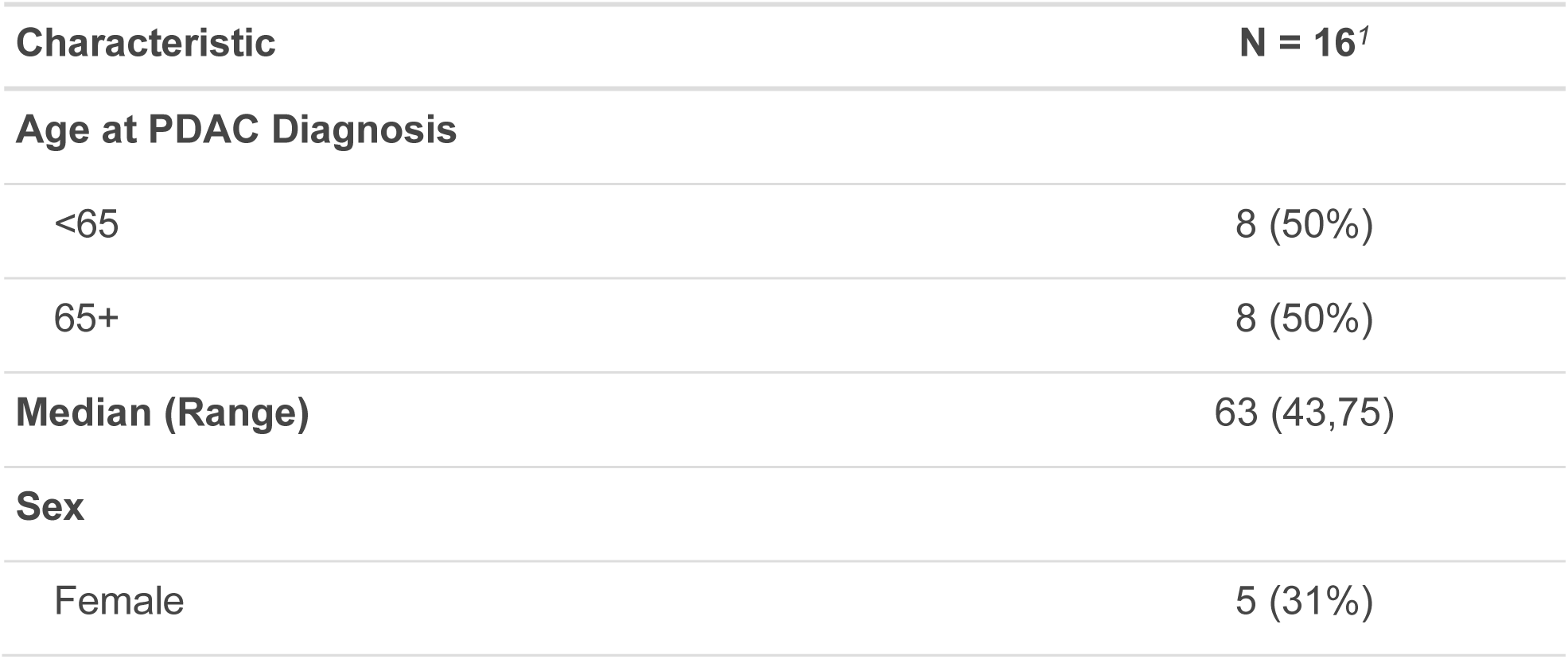
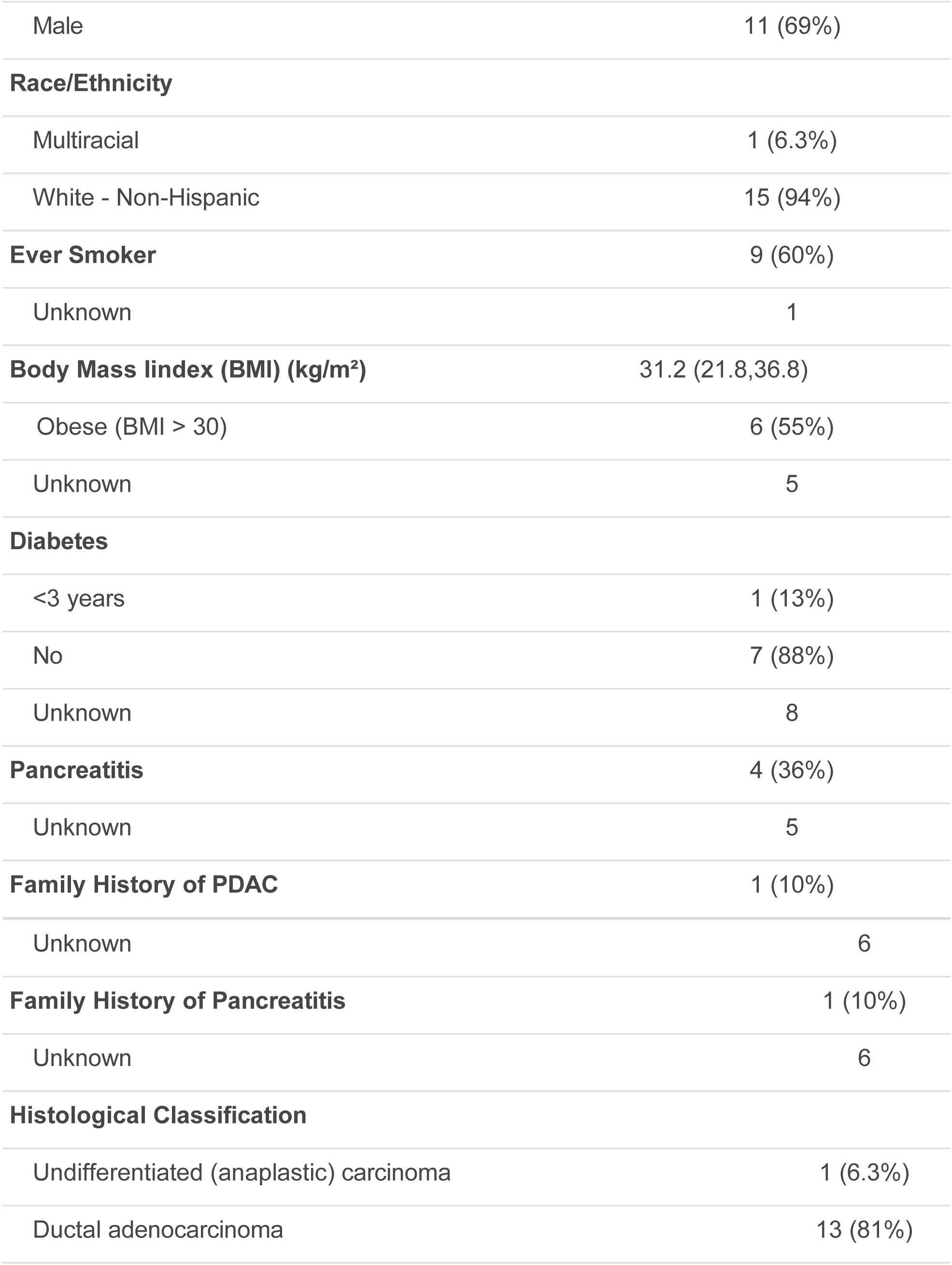
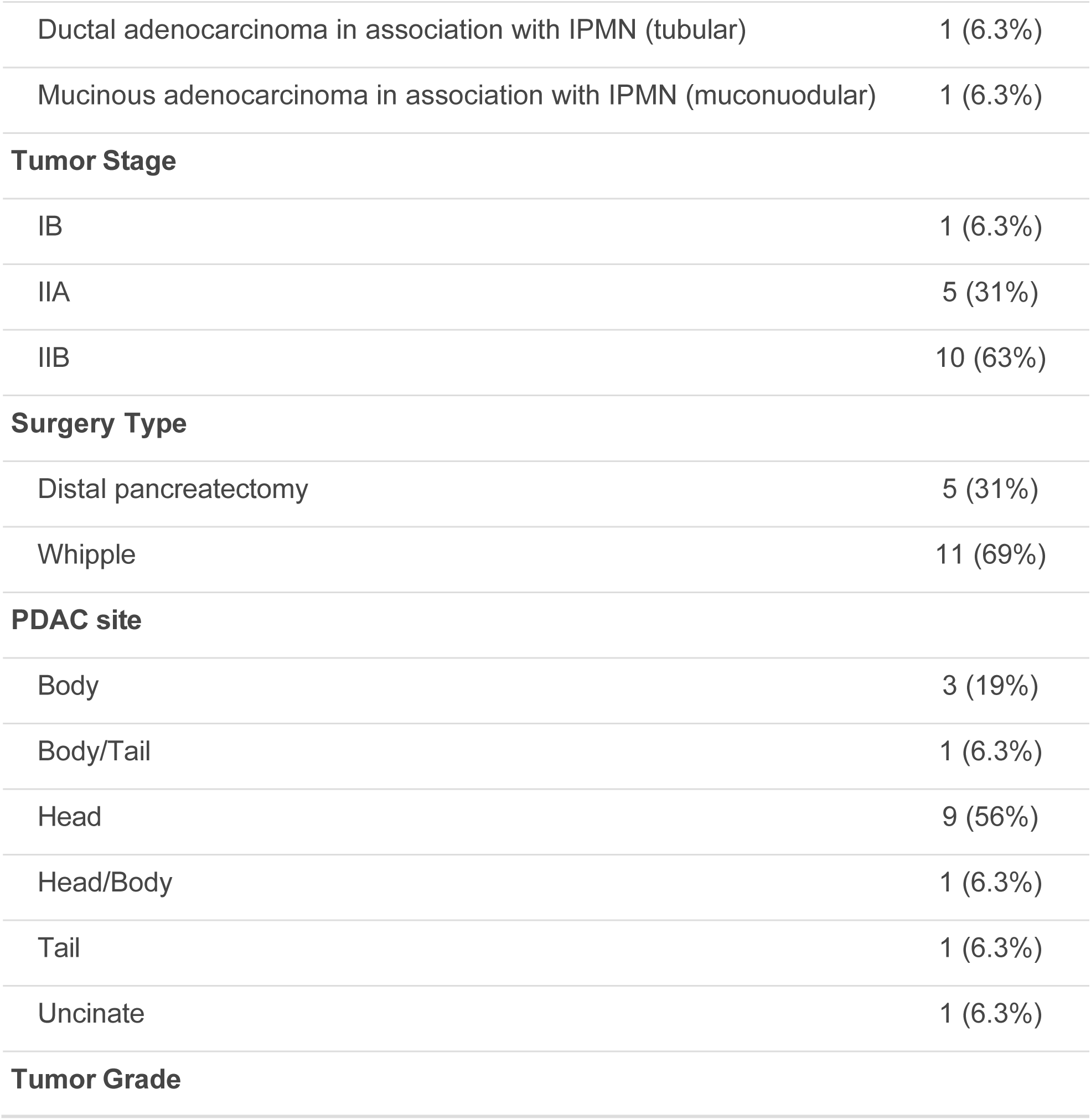
Select patient characteristics for 16 patients with paired samples.

For alignment and quantification of the Mayo RNA-Seq data, we used a pipeline based on HiSat2 and subread (CHURP(Baller et al., 2019), HiSat2(Kim et al., 2019), and featureCounts(Liao et al., 2014)). The TCGA and Mayo data sets were analyzed independently for deconvolution. For each data set, raw counts were normalized to transcripts per million (tpm). Cell type reference signatures were created from two different published single cell RNA-seq studies: sig1 from GSE229413 and sig2 from GSE205049.

Annotations from the original publications were used to define cell types and to subset cells to those from tumor samples (not normal tissue). We filtered each signature set to remove genes with all zeros in all samples and then filtered each data set to remove any genes with mean expression across all samples that was less than the overall median gene expression value. This was done to ensure that the most robust genes were kept for the signatures. We kept only the genes that were found in both scRNA-seq signatures to have the same set of genes. We then used granulator(Pfister S et al., 2024) to run deconvolution for each bulk RNA-seq data set with each scRNA-seq reference signature. The granulator R package is designed to run multiple deconvolution algorithms, and we report the non-negative algorithm results for the methods defined by the granulator package as dtangle, nnls, and qprogwc. We used these three different deconvolution methods to compare cell type proportion estimates for both runs of RNA-seq separately.

We evaluated the impact of batch correction and normalization on median expression. We present in the main manuscript the log2 normalized study-specific gene expression values per gene by calculating tpm across all shared genes and then compare median expression values of key pancreatic cancer genes between studies. We use the ComBat-seq function in the R package sva to adjust for batch effect using an empirical Bayes framework. EdgeR to was used to normalize the batch corrected counts within gene and transformed the values by the log2 function. We selected *KRAS, TP53, SMAD4*, and *CDKN2A*, as these are the genes which are commonly mutated in PDAC.(H. feng Hu et al., 2021) We also selected *CTNNB1, JUN, SMAD3, SMAD7*, and *TCF7*, as these transcription factor enrichment identified genes have been observed to have altered gene expression in PDAC compared with adjacent normal tissue.(Atay, 2020)

Using a reference scRNA-seq study (GSE205049), we evaluated median differences in cell type proportion between the PDAC sample and the adjacent normal paired tissue samples from 9 patients using a Wilcoxon Rank Sum Test for 23 cell types. After deconvolution, we created scatter plots to show the correlation across the two RNA-seq tumor samples for each patient and evaluated significant differences in median cell type proportion using a paired Wilcoxon rank sum test for 21 cell types across the two paired samples. We used the ESTIMATE algorithm implemented in the tidyestimate package in R to estimate tumor purity (stromal and immune cell signatures) using the normalized RNA-seq data from each study separately. Additionally, we evaluated median differences using a paired Wilcoxon rank sum test across the selected nine genes previously determined to be important in PDAC. We visualized differences in log2 normalized gene expression using violin plot and statistically evaluated the significance of the median difference using a Wilcoxon Rank Sum Test with Holm adjusted p-value with values <0.05 indicating significance. We also explored the effects of batch correction, normalization, and transformation on median differences. A plot was also generated to show correlations between each of the genes based on Spearman correlation coefficients.

## Results

We observed that average cell type proportion varied the most between studies (i.e., between samples for each patient) for NK and macrophages (**Figure 1A and B**). We visually observed across all plots that Macrophages, NK cells, T cells, cycling, and dendritic cells are among the cell types that vary the most. Based on the two single cell reference samples, we observed that NK cells are significantly (adj p-value <0.05) enriched in PDAC tissue compared to adjacent normal tissue while Dendric cells (DC1 & 2) and CD16+ monocyte populations are significantly reduced (**Figure 2A**). Using the Wilcoxon test in our paired samples, we observe the most significant difference in the deconvolution-based cell types are for CD4 (adjusted p-value = 0.00613), endothelial (adjusted p-value = 0.00000172), fibroblasts (adjusted p-value = 0.000161), granulocytes (adjusted p-value = 0.000895), neural (adjusted p-value = 0.000378), NK (adjusted p-value = 0.00224), and monocytes (adjusted p-value = 0.00816; **Figure 2B**).

**Figure 1.**
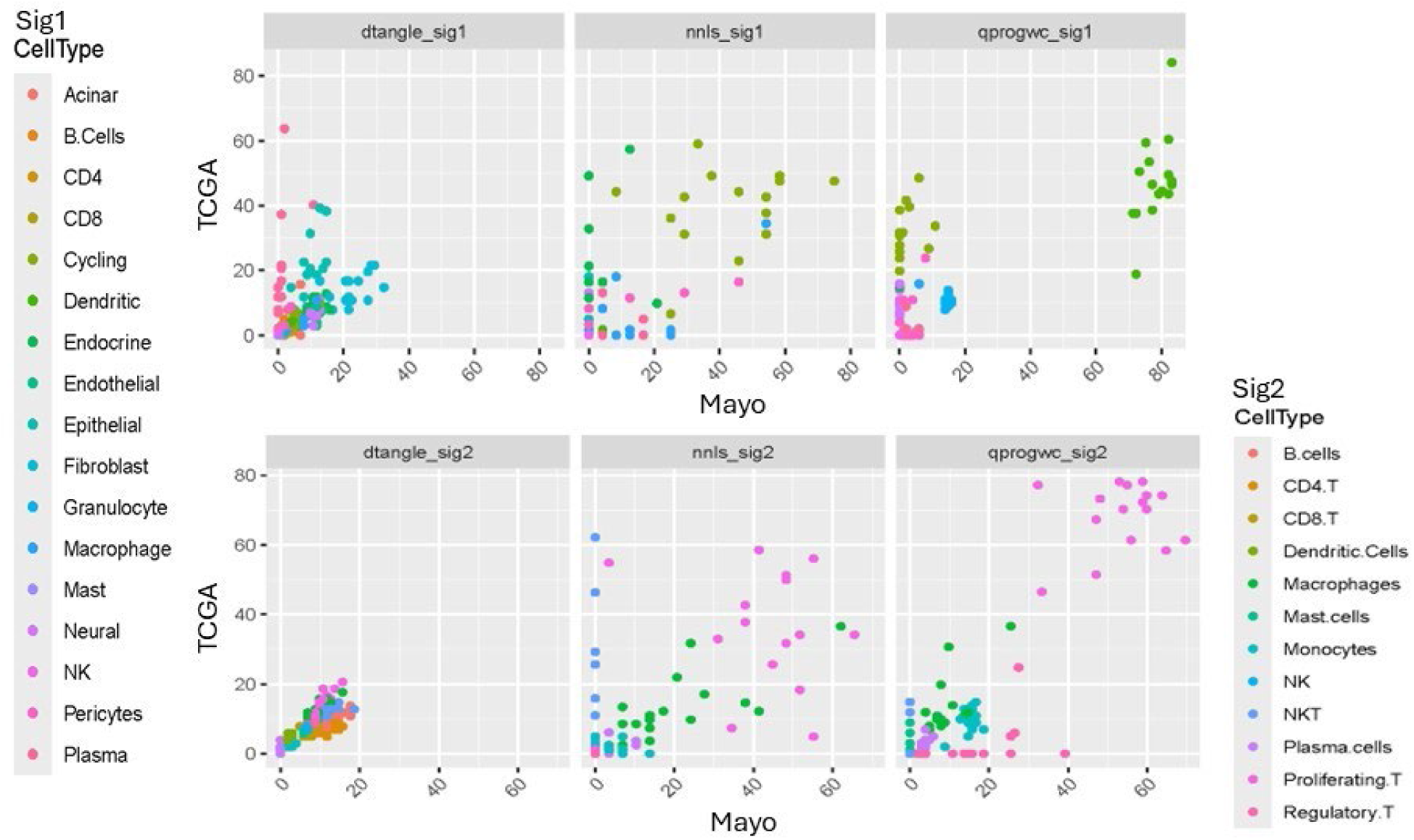
**A)** Sig 1 **& B)** Sig 2 deconvolution cell type proportion estimates for 16 paired samples. Each panel represents a different deconvolution method. Each color represents an individual cell type. Each point represents a patient’s proportion for The Cancer Genome Atlas Program (TCGA) sample (y-axis) and the second study sample (x-axis).

**Figure 2.**
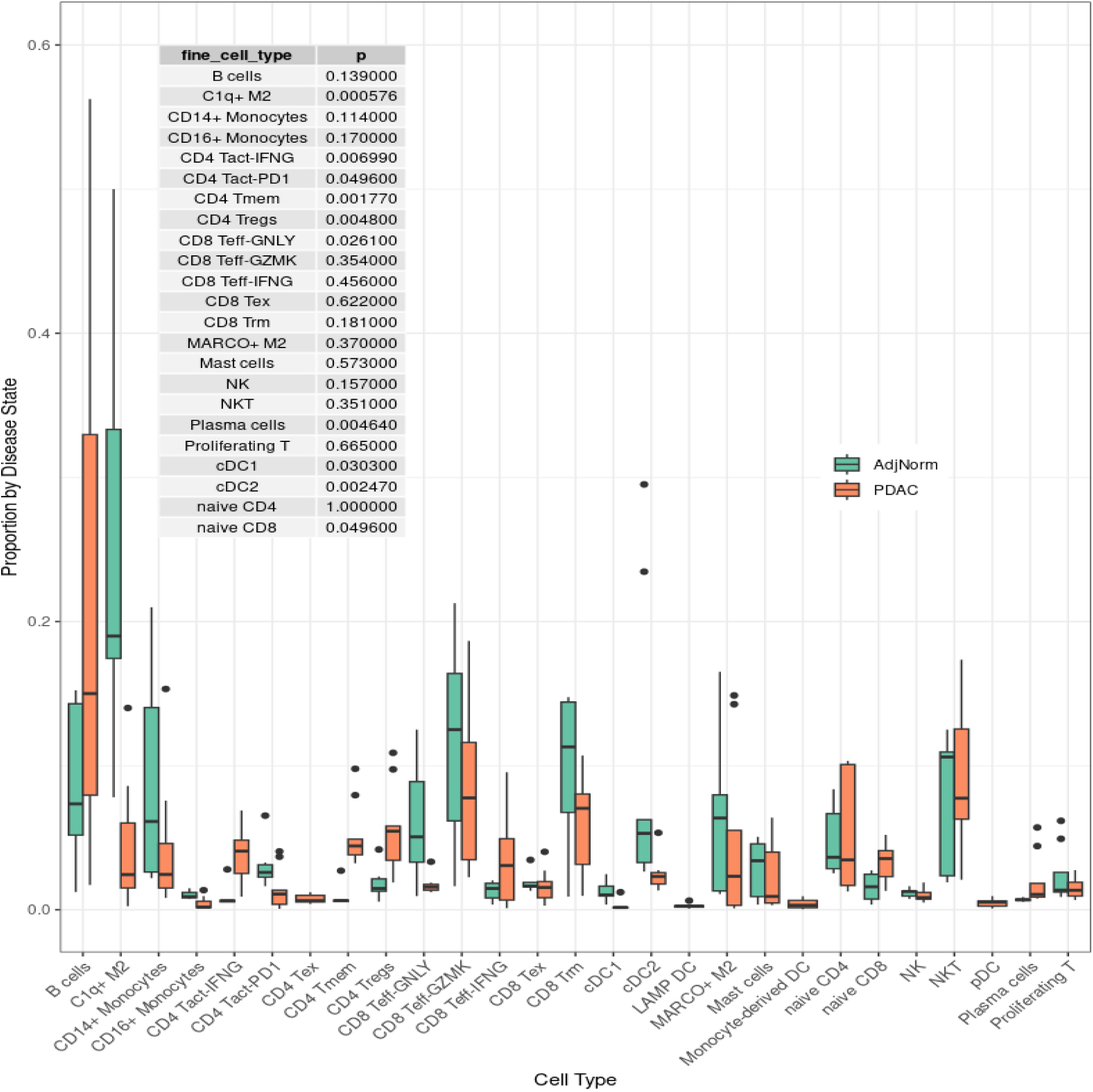
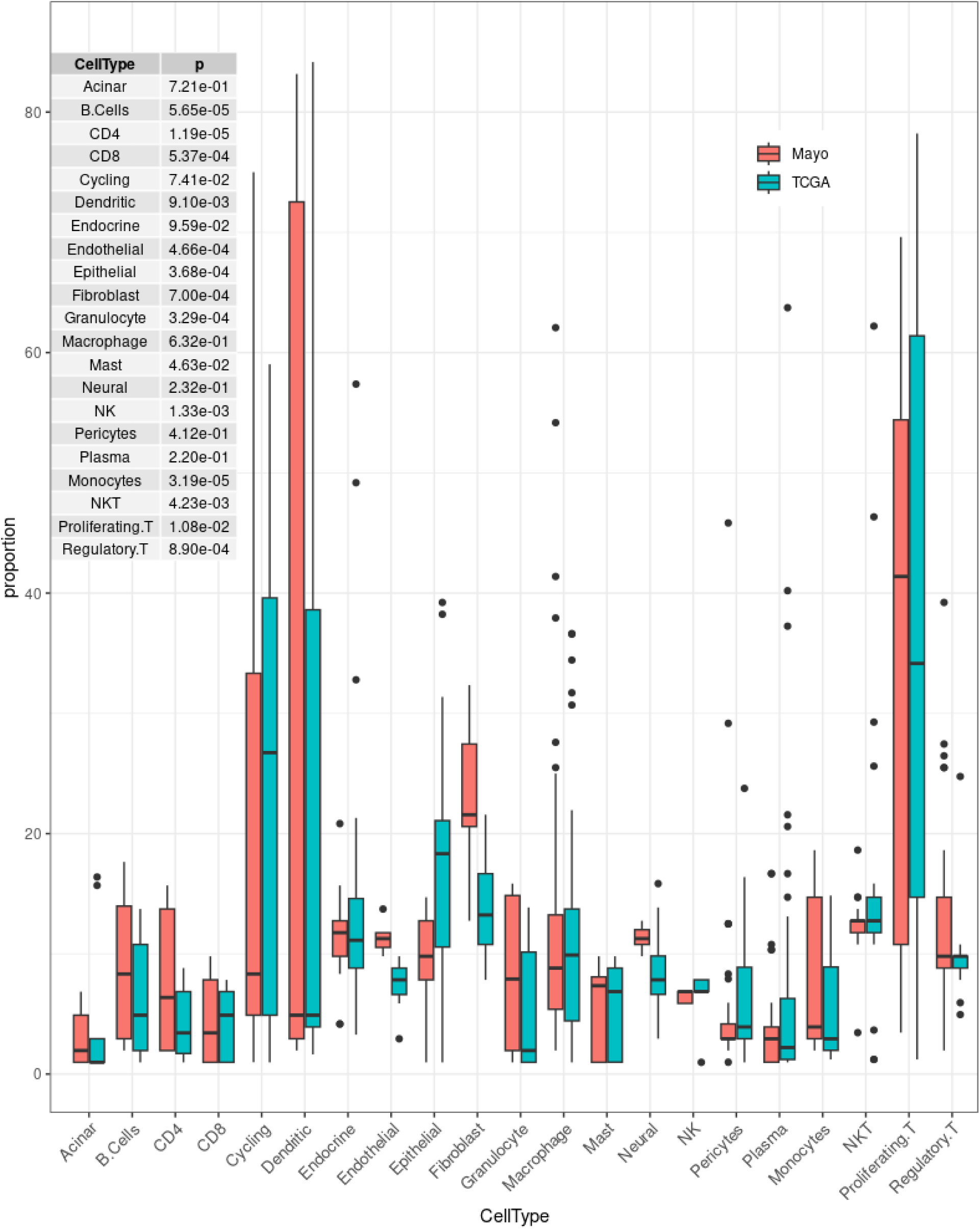
**A)** Comparison of PDAC (orange) and Adjacent normal (green) cell type proportions. **B)** Comparison of Mayo (salmon) vs TCGA (blue-green) cell type proportions across 21 different cell types. P-values are from the Wilcoxon rank sum test. The scatterplot shows the correlation between TCGA RNA-seq-based deconvolution results and Mayo RNA-seq-based deconvolution. The top row represents more cell types and uses a different single cell reference sample than the bottom row. Each of the three columns represent a different deconvolution method: dtangle, nnis, and qprogwc. Each point is colored according to the cell type, as indicated by the legend on the left and right sides. The primary focus should be on identifying patterns for specific cell types that deviate from the diagonal.

We estimated tumor infiltration scores in each study separately using normalized RNA-seq data in the ESTIMATE function in R. In summary, this plot (**Figure 3**) demonstrates the positive relationship between the estimated stromal and immune cell infiltration and the ESTIMATE score, suggesting that tumors with high stromal and immune cells also likely have high infiltration of the tumor. This is expected because a higher presence of non-tumor cells will naturally lower the proportion of tumor cells in the sample. The blue-green (TCGA) and salmon (Mayo) circles highlight specific tumor samples with distinct characteristics in terms of their microenvironment. It is important to note that ESTIMATE scores can only be interpreted relatively, and it cannot be inferred across studies.

**Figure 3.**
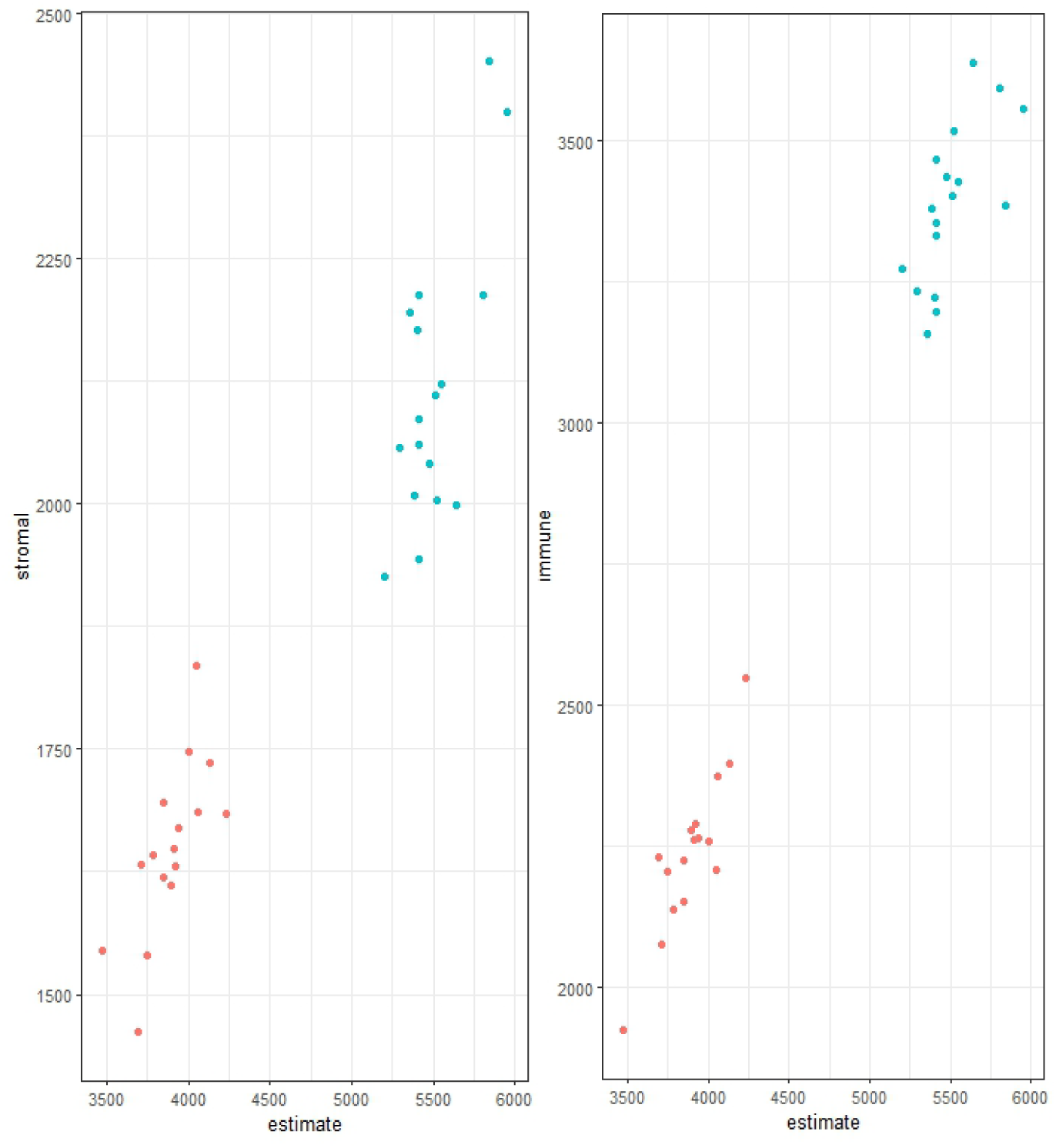
Tumor infiltration estimates for Mayo (salmon) and TCGA (blue-green) sample data. The ESTIMATE algorithm from R was used to estimate A) stromal and B) immune infiltration in the sample using the normalized gene expression data for each study separately. The tumor infiltration estimate is relative and cannot be compared across studies.

Violin plots of gene expression in the paired Mayo and TCGA sample data for the four PDAC specific genes comparing the effect of batch correction, normalization, and transformations are included (**Supplemental Figure S1a and b**). The plots highlight the importance of adjusting both within sample (normalization) and across sample (batch correction) to avoid bias. When looking at the first four PDAC specific genes, we did not observe significant median expression differences across paired samples. However, there may be hints of differences for KRAS (adj. p-value = 0.18) and SMAD4 (adj. p-value = 0.59, **Figure 4A**). When evaluating the five transcription factor enriched genes, there were no statistically significant results. Visually, CTNNB1(adj. p-value = 0.30) and TCF7 (adj. p-value = 0.56) varied the most between the paired TCGA and Mayo sample data (**Figure 4B**).

**Figure 4.**
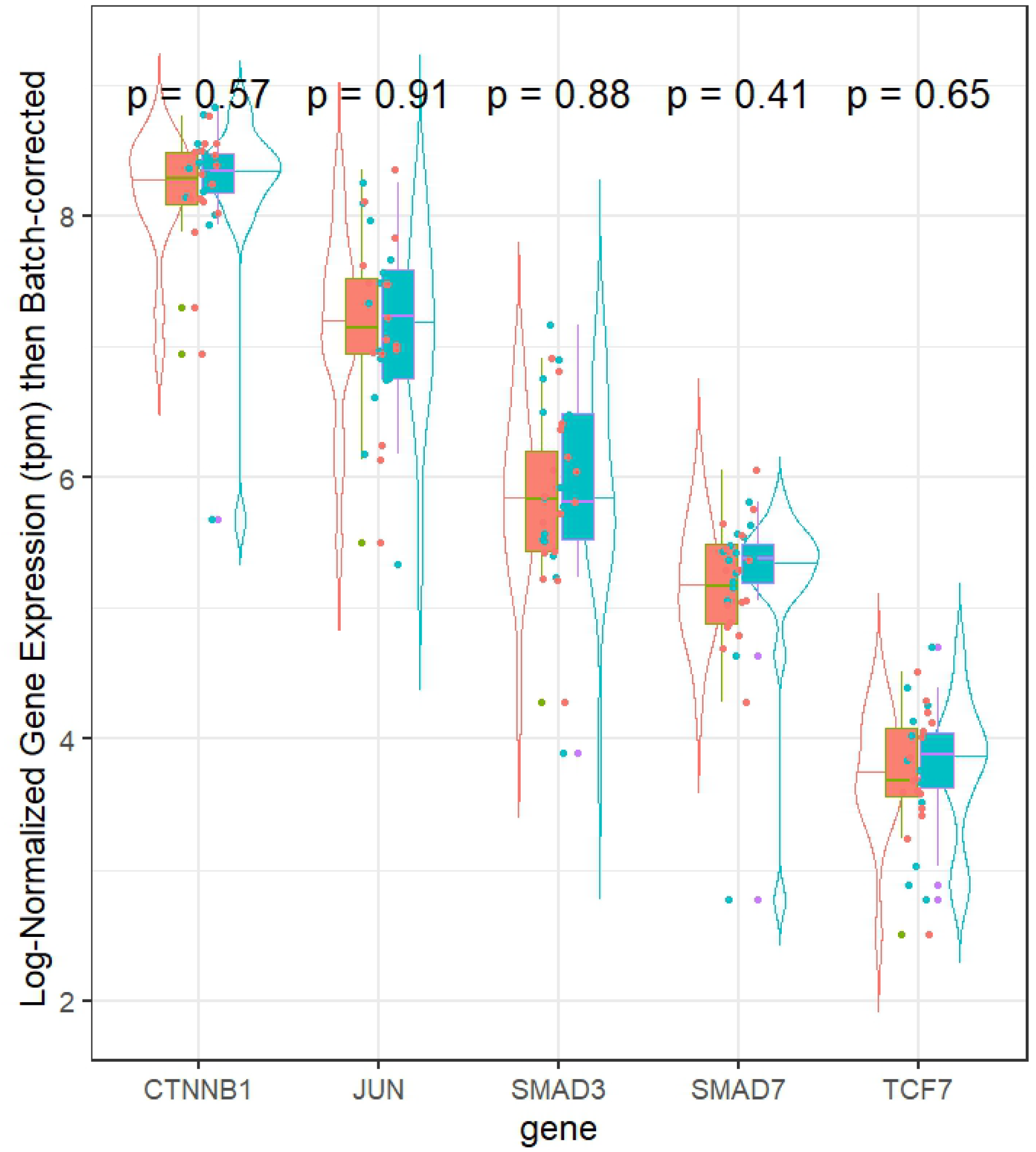
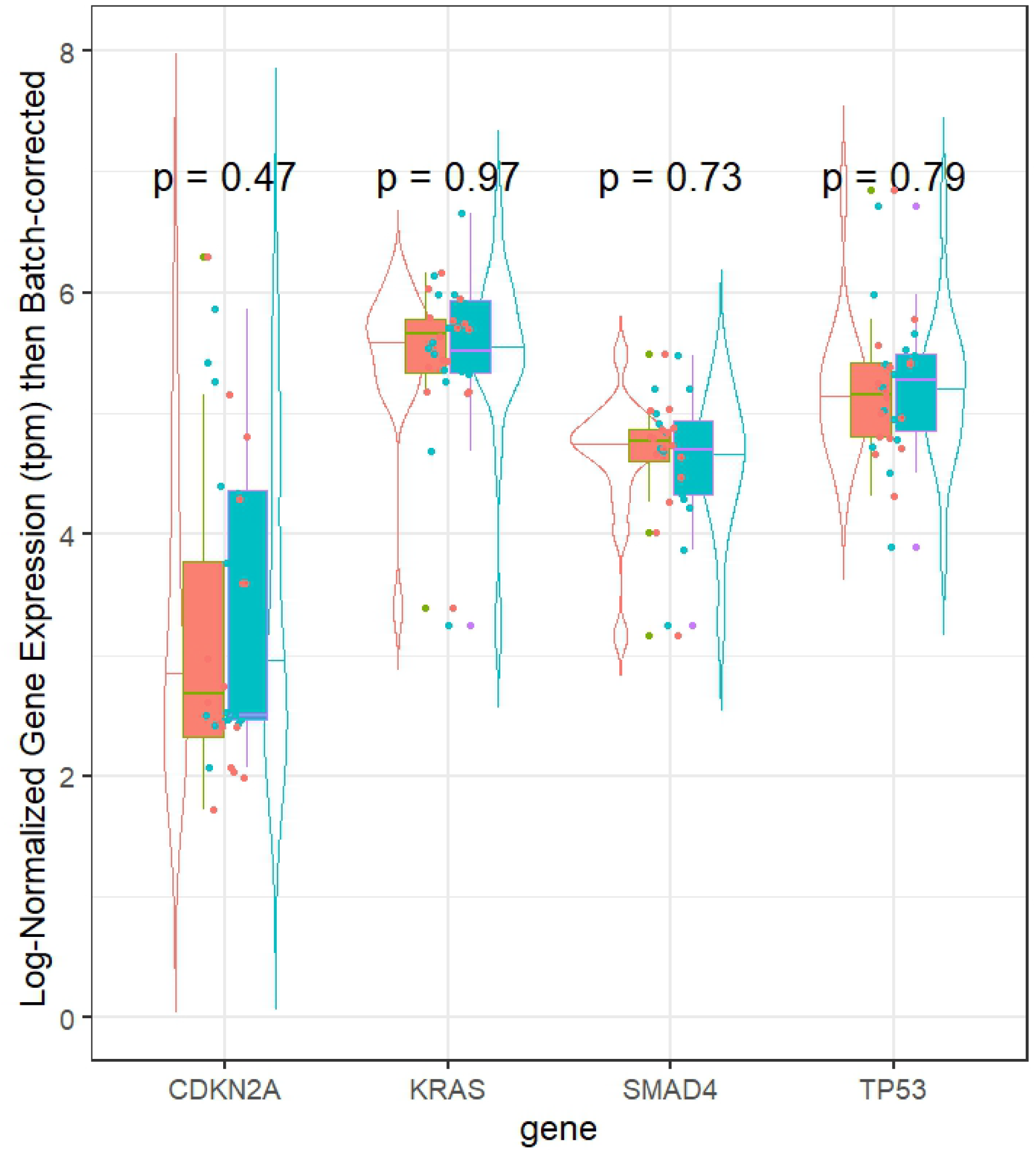
Violin plot of normalized gene expression in transcripts per million (tpm) for Mayo (salmon) and TCGA (blue-green) samples in **A)** four selected highly mutated pancreatic cancer genes and **B)** five validated highly variably expressed genes. Each gene also has an adjusted p-value for the Wilcoxon comparison between paired samples.

We used the median as a cut point for each gene and calculated one score for each patient separately to determine concordance. (**Table S1**) We observed a substantial concordance (kappa=0.75) for *JUN*, moderate concordance for *KRAS, TP53, SMAD3* and *CDKN2A* (Kappa =0.5) and low concordance value for *TCF7*, *SMAD7, SMAD4, CTNNB1*(Kappa = 0.25) across the two studies suggesting that sufficiently representative genomic data cannot be collected with one sample. Likewise, the correlation of the gene expression values for the nine selected genes in general vary significantly across the paired samples (**Figure 5)**. In this correlation matrix, each row and column represent a gene and the number in the cell represents the Spearman correlation coefficient with colors ranging from highly positively correlated values (blue) to highly negatively correlated values (red). The coefficients which reached statistical significance ranged from <-0.24 to >0.24 and are shown in the upper triangle. The hierarchical clustering method was used to order the genes in the matrix. We observed moderate positive correlations between TP53/SMAD4 (0.47) among only TCGA samples; SMAD7/CTNNB1 (0.69), KRAS/CTNNB1 (0.36), and SMAD4/CTNNB1 (0.31) using only Mayo samples. We observed a moderate negative correlation between TCF7/CDKN2A (-0.24) within the TCGA samples only. We observed moderate positive correlation across study genes for CDKN2A/CDKN2A (0.52), SMAD4/TCF7 (0.47), and SMAD3/CDKN2A (0.40). Moderate negative correlations were observed for the following genes across studies: SMAD4/CDKN2A (-0.64), TCF7/SMAD3 (-0.46), SMAD7/TP53 (-0.31), and TCF7/CDKN2A (-0.24).

**Figure 5.**
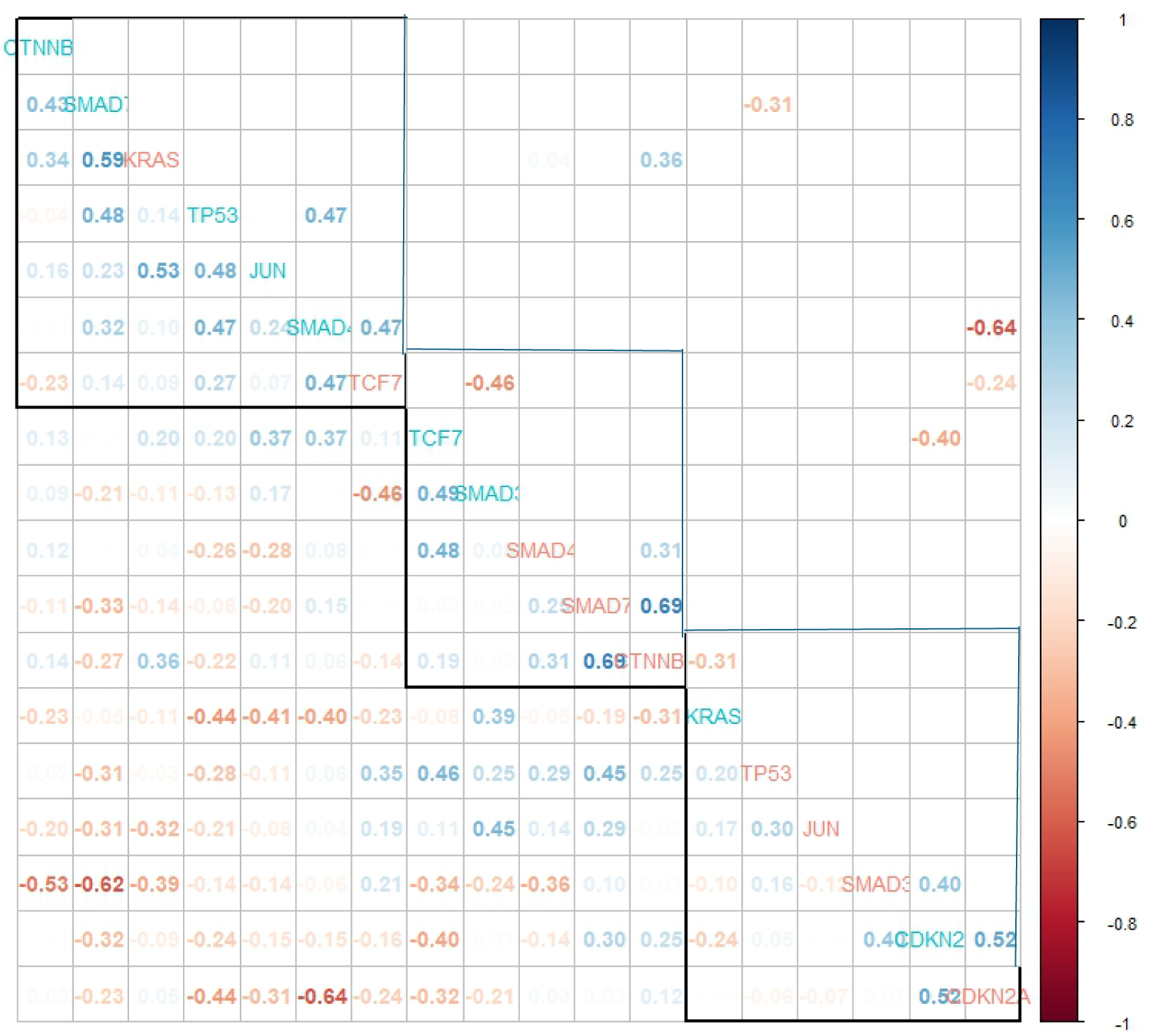
Correlation plot showing the Spearman correlation in select gene expression values for TCGA data (blue-green) and Mayo data (salmon). The genes are arranged based on hierarchical clustering. The lower triangle shows all correlation coefficients with higher correlations having higher transparency. The higher triangle shows only the correlation values which have significant p-values. Correlation coefficients with values close to 1 have deeper blue and coefficients with values close to -1 have deeper red color.

## Discussion

In this study, we observed minimal nonsignificant differences in expression values for selected genes established to have a role in PDAC. This slight difference could be related to the different cell type proportions across the samples that we estimated using deconvolution methods. Studies show that pancreatic tumors are heterogeneous and different regions of the tumor may have distinct genetic and molecular profiles, (Lodestijn et al., 2021) so observing a difference across samples is not surprising. We did not observe significant differences in median log normalized, batch-corrected expression values for our selected genes. However, this may be due to a small sample size. Collectively, the results suggest there should be additional research investigating how closely a single sample fully captures the heterogeneity of a patient’s tumor and determine how important this might be for treatment planning as genomic results are increasingly used in a clinical setting. Focusing research on how many samples to obtain and strategically deciding where to biopsy will be important as single cell and spatial technologies advance.

When evaluating median gene expression across studies, we did not observe any statistically significant differences; however, there may be hints of a difference. Performing follow-up scRNA-seq studies with a larger sample size could help clarify if there is an actual difference by looking at cell type specific expression. As we see using the deconvolution and tumor infiltration estimates, there is large variation in the estimated cell type proportions and tumor cells in the sample.

The correlation matrix heatmap provides a comprehensive overview of the pairwise relationships between the expression levels of the given genes. It highlights strong positive and negative correlations that can provide valuable insights into gene co-regulation, functional relationships, and potential regulatory mechanisms. We used hierarchical clustering to order the genes, and that method grouped the genes with sample type intermingled (Mayo vs. TCGA). This could indicate that because cell type proportions vary across samples, there are also potential co-regulation pathways that differ by cell type. Additionally, the difference in correlation structure by sample type could suggest a difference in functional relationships or regulatory mechanisms utilized across the two samples. Cross-sample correlations could potentially suggest a regional relationship between the genes or could just represent random association. Mechanistic studies in the laboratory would need to confirm any of these suggested relationships.

Some of the key limitations in this research include the small sample size and that each of the paired samples were processed using different sequencing technologies. Any findings should be confirmed with additional studies.

The consistency of RNA-seq results across multiple samples from the same person is a complex issue. Factors that can influence consistency include: 1) biological variation as gene expression fluctuates due to circadian rhythms, physiological state (e.g., stress, illness), and cellular heterogeneity and 2) technical variation as there are many processing steps in the RNA-seq workflow and bioinformatic analysis.(Auer & Doerge, 2010) Careful experimental design and quality control are essential to minimize technical variation. While some variability is inherent in biological systems, RNA-seq can provide highly consistent results when proper experimental design and analysis are employed. (Su et al., 2014)

Tumor heterogeneity between different sites of neoplasia in a single patient, and among tumors from different patients, is an emerging theme in cancer research.(Alizadeh et al., 2015) Heterogeneity confounds researchers’ understanding of tumor evolution and their ability to design effective treatments.(Javed et al., 2024; Y. Liu et al., 2024, 2025) Due to the limitations of methods that characterize tumors in ‘bulk’, such as averaging across all tumor clones, even a single RNA-Seq sample, may not fully represent the tumor heterogeneity.(Y. Liu et al., 2024, 2025) Ideally, researchers would obtain multiple regions from each tumor to capture spatial heterogeneity.(Giannoukakos et al., 2024; Sergeyev et al., 2024; Yaqoob et al., 2024)

While computational deconvolution of different expression components in a sample can distinguish between cells from different lineages, it has limited applicability in samples with low transcriptional diversity.(Y. Liu et al., 2025) Even at the single cell level, several factors affect the ability of a single RNA-seq sample to represent tumor heterogeneity.(Hwang et al., 2022) Single-cell RNA-Seq is a robust technology that can analyze tens of thousands of cells simultaneously in a cost-effective and efficient manner.(Hwang et al., 2022) However, sensitivity for low expressed genes still presents a challenge, and metrics that quantify similarity and difference between samples from the same clonal origin are needed.(Hwang et al., 2022; Y. Liu et al., 2025)

The long-term goal of this area of research is to harness the power of genomic analysis to optimize biopsy decisions and ultimately improve patient outcomes. Challenges remain in fully validating genetic and genomic biopsy strategies and supporting their widespread clinical implementation and will be the focus of future work. Attention should focus on standardizing tissue collection/processing protocols, establishing evidence-based thresholds for acting on genomic findings, and integrating molecular and image data into existing clinicopathologic risk assessment frameworks. Research in the future should focus building off the results presented here to get a clearer picture if multiple samples taken at the time of biopsy would help develop more effective treatment plans.

## Data Availability

All data produced in the present study are available upon reasonable request to the authors.

https://portal.gdc.cancer.gov/

