## Supplemental Figure S1a-b, Supplemental Table S1 for "Variation in bulk RNA-seq and estimated cell type proportion using deconvolution when comparing pancreatic cancer samples within the same individual"

**Supplemental Files**

**Figure S1a**. Comparisons of “normalized”, “batch corrected (no normalization)”, “batch and normalized”, and “batch and log2 normalized” gene expression of Mayo (left) and TCGA (right) paired samples for four PDAC specific genes.

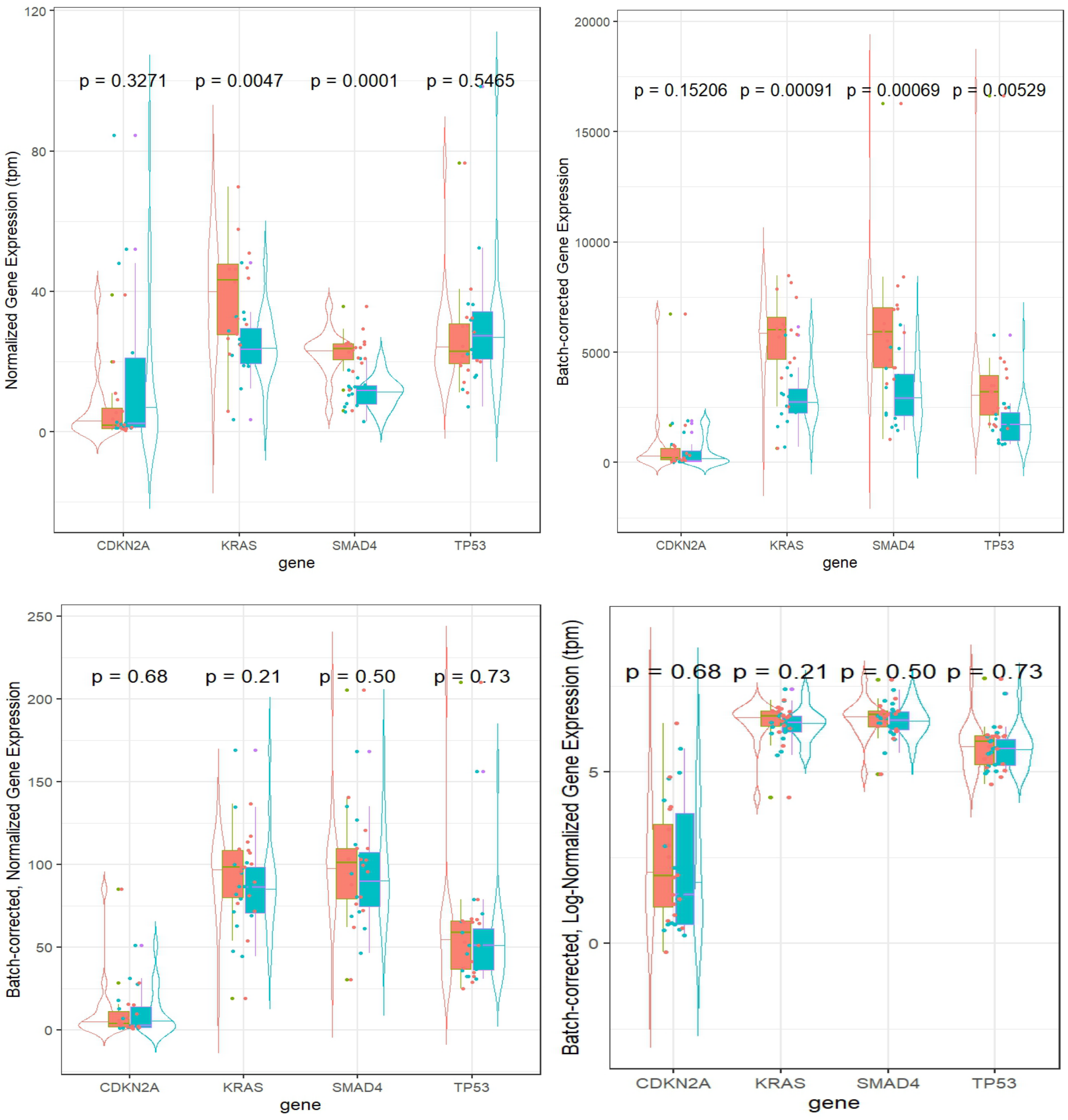

**Figure S1b**. Comparisons of “normalized”, “batch corrected (no normalization)”, “batch and normalized”, “batch and log2 normalized” gene expression of Mayo (left) and TCGA (right) paired samples for five genes commonly mutated in PDAC.

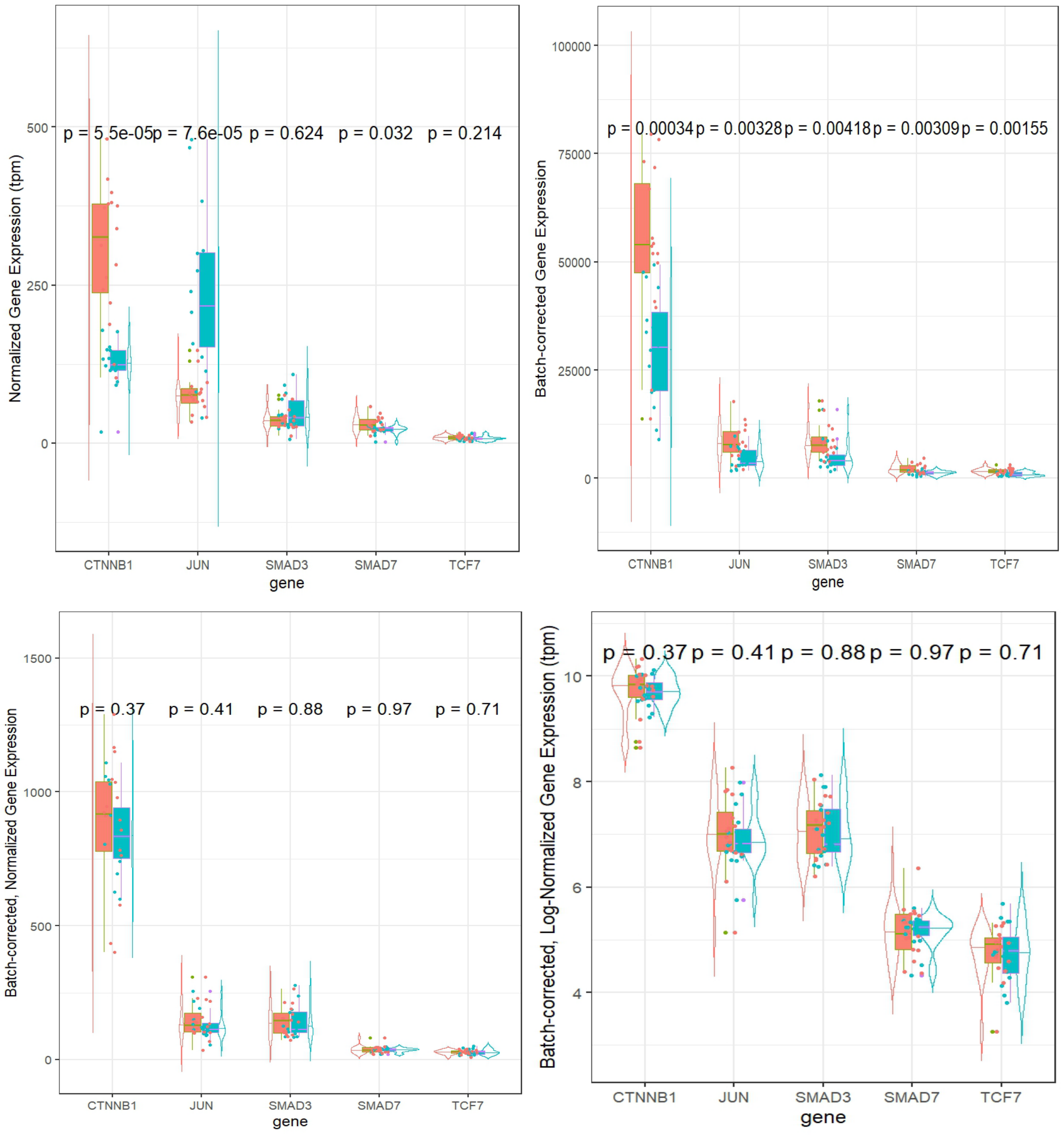

**Table S1**. Kappa statistic and p-value across selected genes

| Gene | Kappa | p-value |
|---|---|---|
| *KRAS* | 0.5 | 0.0455 |
| *TCF7* | 0.25 | 0.317 |
| *SMAD7* | 0.25 | 0.317 |
| *TP53* | 0.5 | 0.0455 |
| *SMAD4* | 0.25 | 0.317 |
| *SMAD3* | 0.5 | 0.0455 |
| *CTNNB1* | 0.25 | 0.317 |
| *JUN* | 0.75 | 0.0027 |
| *CDKN2A* | 0. 5 | 0.0455 |
